# The Impact of Heads Up Testing on Thrombectomy for Acute Ischemic Stroke

**DOI:** 10.1101/2025.03.08.25323604

**Authors:** Mallory Blackwood, Charles Beaman, Latisha Sharma, David S. Liebeskind

**Author notes:** Corresponding author: Mallory Blackwood, 710 Westwood Plaza, Los Angeles, CA 90095.

## Abstract

The Heads Up test, initially described in 2017, offers a potential tool for assessing candidacy for mechanical thrombectomy (MT) in patients with acute large vessel occlusion (LVO) but minimal neurological deficits. This study aims to examine the practical applications and outcomes of the Heads Up test over nine years by analyzing 15 consecutive cases of documented Heads Up testing. By assessing clinical outcomes, interventions, and the demographic factors influencing decision-making in Heads Up-positive cases. Our findings suggest that the Heads Up test can provide valuable guidance in treatment decisions, but further data is needed to refine its criteria and applicability in the evolving neurointerventional practice.

## Introduction

Since its inception, mechanical thrombectomy (MT) has revolutionized acute ischemic stroke treatment, especially for large vessel occlusions (LVOs) (Lambrinos et al., 2016). The Heads Up test, introduced in the literature in 2017, was developed to support decision-making for MT in cases where patients present with LVO but exhibit only mild neurological symptoms (Ali et al., 2016). By elevating the head-of-bed to 90 degrees for a duration of 30 minutes and assessing for change in the NIH Stroke Scale (NIHSS), the Heads Up test aims to identify patients at risk of functional deterioration. A positive Heads Up test is defined as any worsening of neurological status during the test period, prompting its termination and consideration for MT.

Since the initial description of Heads Up testing, the landscape of neurointerventional has evolved, with broader and more nuanced criteria for MT eligibility (Jadhav et al., 2021; Pfaff et al., 2016). Here, we present a retrospective analysis of real-world Heads Up test use between 2015 and 2024, discussing the decision criteria, procedural outcomes, and complications in 15 cases to further elucidate the clinical role of Heads Up testing in determining MT candidacy.

## Methods

This study analyzed 15 patients with acute LVO who underwent Heads Up testing between 2015 and 2024. Charts were reviewed to acquire patient demographics, NIHSS scores, Heads Up testing rationale, test outcomes, and treatment decisions. Patient comorbidities, prior antiplatelet or anticoagulation use, and details of intervention were also reviewed.

The Heads Up test was performed by elevating the patient’s head to 90 degrees and monitoring for any NIHSS changes over 30 minutes. The test was considered positive if any deterioration occurred, prompting consideration for MT. Interventions, including MT techniques (e.g., aspiration, stent retrievers) and adjunctive procedures (e.g., angioplasty, stenting), were recorded. Patient outcomes, including eTICI scores, symptomatic intracranial hemorrhage (ICH, defined as any clinical worsening with PH1 or greater bleed) and NIHSS scores post-intervention, were analyzed.

## Results

Identifying Heads Up cases proved more difficult than anticipated due to a lack of standardized documentation. Cases were identified by a search of the EHR, and only cases where Heads Up testing in the IR suite was mentioned, with documented exam were included in the assessment.

Of the 15 patients who underwent Heads Up testing, six demonstrated worsening neurological function and were classified as Heads Up-positive. Nine patients did not demonstrate neurological worsening and were classified as Heads Up-negative. MT was performed in eight cases during the hospital stay; six cases were conducted for positive Heads Up testing, and two cases were initially Heads Up-negative but required delayed intervention due to clinical worsening. The negative predictive value of the Heads Up test in our series was 82%. All eight MT cases achieved eTICI 2b or higher, indicating successful revascularization.

A combined approach of aspiration and stent retrievers was most common, with six of eight cases achieving favorable outcomes, while aspiration alone was effective in only two cases. Two patients required permanent stent placement, and one required angioplasty for significant intracranial stenosis. Symptomatic ICH occurred in two cases: one who received MT and one who did not, both of whom had initially negative Heads Up testing All 15 patients had anterior circulation occlusion, 3 (20%) had stenosis of the affected vessel. Patient demographics (summarized in table 1) revealed a high prevalence of cardiovascular risk factors: 53% had hypertension, 40% had hyperlipidemia, and 33% had malignancy or hypercoagulability. Notably, eight patients were on antiplatelet or anticoagulant therapy before admission. Other comorbidities included coronary artery disease (20%), atrial fibrillation (20%), prior stroke (6.6%), and diabetes (6.6%).

**Table 1:** Demographics.

| Demographics | Total<br>(N=15) |
| --- | --- |
| Previous antiplatelet/anticoagulation | 8 (53%) |
| Hypertension | 8 (53%) |
| Hyperlipidemia | 6 (40%) |
| Malignancy/Hypercoagulability | 5 (33%) |
| Prior Stroke | 1 (6.6%) |
| ICAD | 3 (20%) |
| Atrial Fibrillation | 3 (20%) |
| Diabetes | 1 (6.6%) |

## Discussion

Our case series offers insights into the clinical utility of Heads Up testing for assessing MT candidacy in patients with acute LVO and low NIHSS scores. Heads Up testing was most frequently employed in cases where there was uncertainty regarding the need for intervention, such as low or resolving NIHSS scores or imaging suggestive of proximal occlusion despite minimal symptoms.

Our findings suggest that approximately half of Heads Up-tested patients required MT during hospitalization, with two cases initially deemed Heads Up-negative later requiring intervention due to neurological worsening. The Heads Up test in our small series demonstrated a negative predictive value of 82% indicating that it may be helpful to rule out the need for MT in certain patients. This highlights the Heads Up test’s potential, and even the intuition to utilize it, though further studies are needed to refine criteria for Heads Up-positive status.

The need for adjunctive procedures (e.g., stenting, angioplasty) in several cases indicates that patients with Heads Up-positive tests may have more complex vascular pathology, potentially predisposing them to re-occlusion. Moreover, the observed rate of symptomatic ICH underscores the importance of cautious patient selection, particularly in those with existing comorbidities, anticoagulation therapy, or delays in decision to intervene.

This study emphasizes the evolving criteria for MT and suggests that Heads Up testing may offer a valuable decision-making tool, especially as inclusion criteria for MT expand. An evolving NIHSS is critical to mechanical thrombectomy decision-making. To do this, positional changes may need to be evaluated with a standard protocol as we have defined with Heads Up. To allow for better quantification of the decision to pursue mechanical thrombectomy in real practice, we recommend that interventionalists familiarize themselves with the Heads Up test and document it in their exam when applicable. Given the limited sample size and observational nature of this study, further research is warranted to validate Heads Up testing as a reliable predictor of MT benefit. Future studies should aim to establish standardized protocols for Heads Up testing and evaluate its predictive value in a diverse patient population.

## Limitations

While some conclusions may be drawn from this retrospective case series, there are of course limitations. The sample size is smaller than expected, in large part due to insufficient documentation. In addition, the retrospective nature carries its own limitations, as we can only guess as to why certain patients underwent Heads Up testing while others did not. We can also not assess the “false positives” or patients who underwent thrombectomy that may not have required it were they subjected to a Heads Up test.

## Conclusion

In this case series, Heads Up testing served as a useful adjunct in the decision-making process for MT in acute LVO patients with minimal deficits. Approximately half of Heads Up-tested patients ultimately required MT, with successful outcomes in cases achieving TICI 2b or higher. The Heads Up test shows promise for identifying patients at risk of deterioration and potential benefit from MT; however, larger studies are necessary to optimize its role in neurointerventional protocols. Further research should focus on refining Heads Up test criteria and integrating it with evolving MT eligibility guidelines to enhance patient outcomes in acute ischemic stroke.

## Data Availability

Data will be provided upon request due to the PHI-sensitive nature of this dataset

## Data Availability

All of Us controlled tier data is available for registered institutions and cohort selection is detailed in the workbench titled "Detecting the prevalence of ultra-rare gene mutations." Due to stringent privacy protections and data sharing restrictions enforced by the All of Us Research Program, individual-level variant data cannot be shared publicly. Researchers must adhere to specific guidelines to protect participant confidentiality, which includes not reporting or disseminating any data that could allow the re-identification of participants, particularly those involving participant counts between 1 and 20 without employing approved data obscuration methods.

## Abbreviations

MT: (mechanical Thrombectomy)
LVO: (Large Vessel Occlusion)
NIHSS: (National Institutes of Health Stroke Scale)

## Acknowledgements/Sources of Funding/Disclosures

None

---

**Figure 1.**
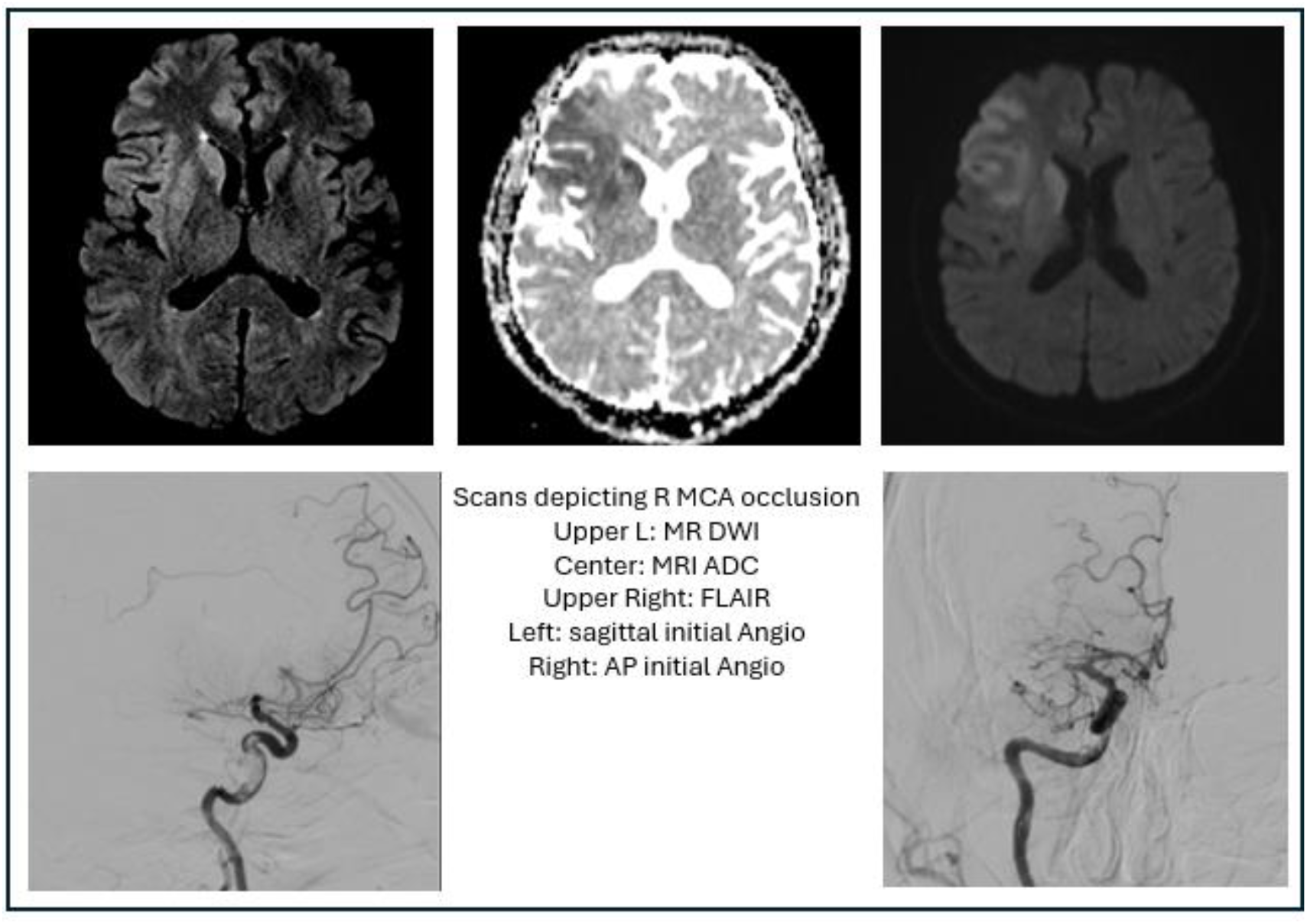
Case 1: Initial NIHSS of 7 with improvement following tPA. Heads up in the IR suite revealed no further worsening, so MT was deferred. Three days later, acutely worsened with NIH 10. MT performed with TICI 2C. Patient required decompressive hemicraniectomy and discharged with trach/PEG.

**Figure 2.**
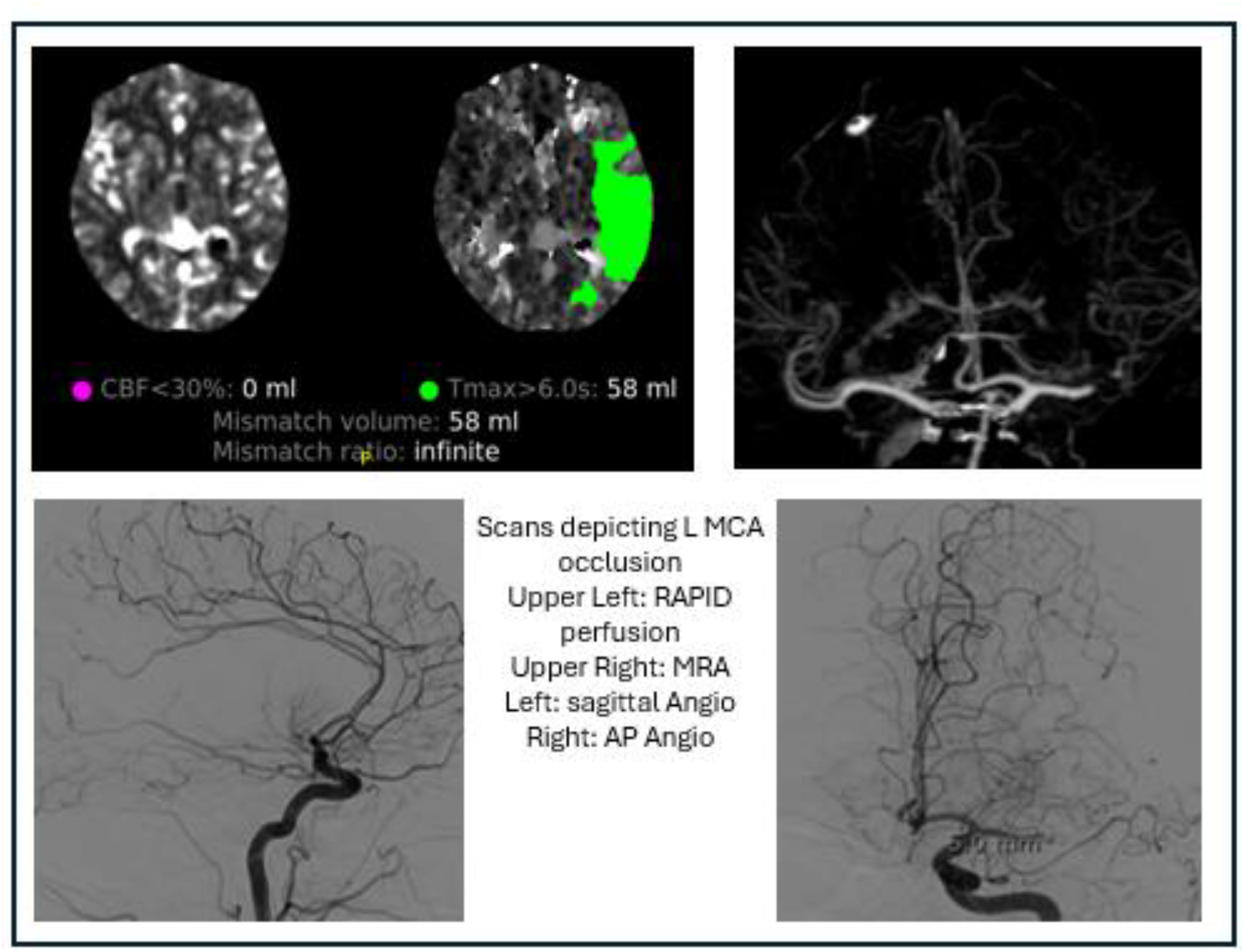
Case 2: Patient presented with wake up stroke and NIHSS 6. A heads up test in IR suite was positive. MT performed with TICI2C and patient was discharged with minimal new deficit.

**Figure 3.**
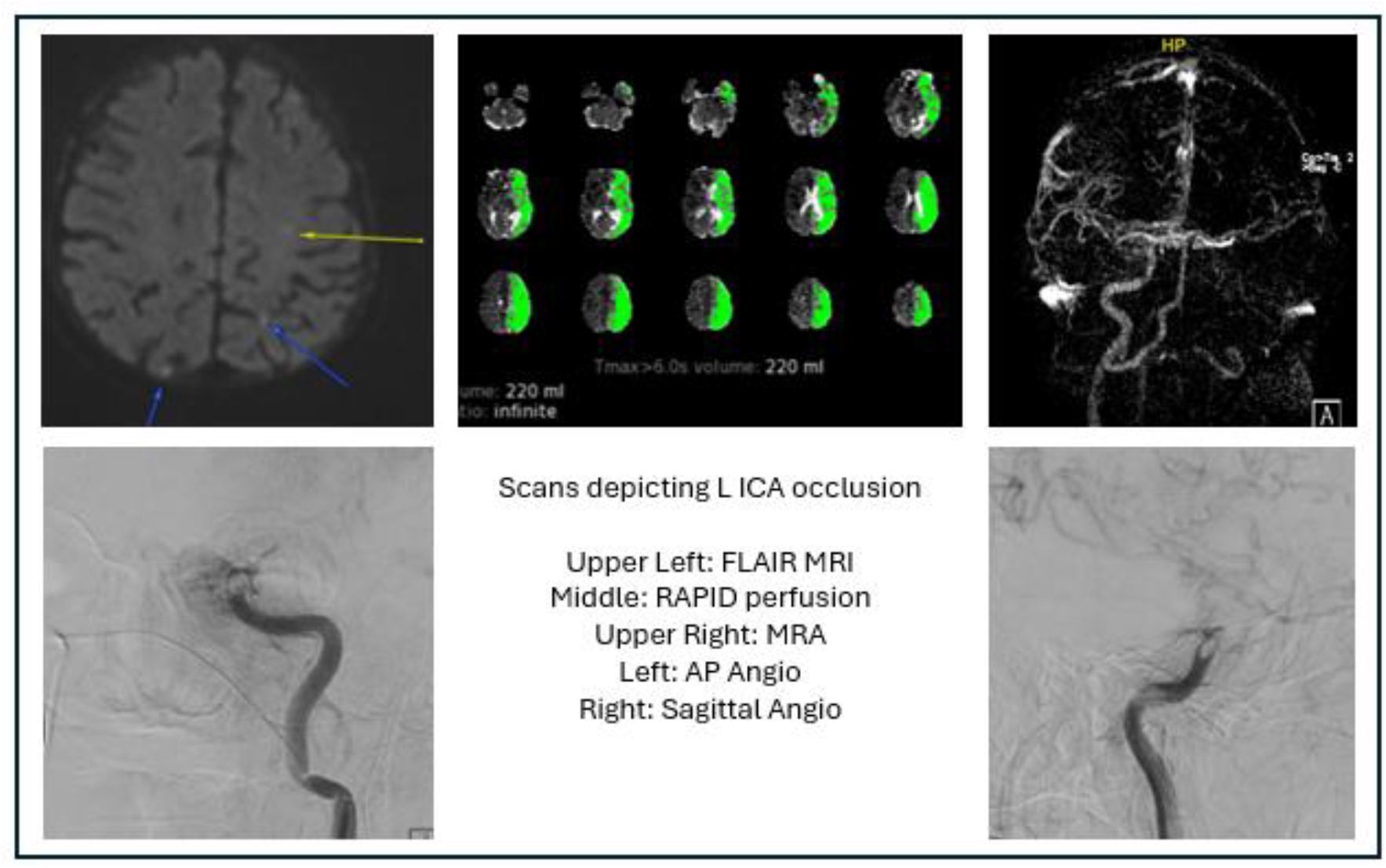
Case 3: Patient with L ICA occlusion, but symptoms improved during transit to IR suite. Heads up done without re-emergence of symptoms. MT not performed, patient stayed in NeuroICU for BP management. MRI day after showed small watershed territory perfusion delay improved with small residual infarct.

